# Comparison of RT-qPCR and Digital PCR Methods for Wastewater-Based Testing of SARS-CoV-2

**DOI:** 10.1101/2022.06.15.22276459

**Authors:** Adrian Hinkle, Hannah D. Greenwald, Matthew Metzger, Melissa Thornton, Lauren C. Kennedy, Kristin Loomis, Monica B Herrera, Raymond-John Abayan, Kara L. Nelson, Rose S. Kantor

## Abstract

Wastewater-based epidemiology is an important tool for monitoring SARS-CoV-2 and other molecular targets in populations, using wastewater as a pooled sample. We compared the sensitivity, susceptibility to inhibition, and quantification of reverse transcription quantitative PCR (RT-qPCR), microfluidic well digital RT-PCR (RT-dPCR), and droplet digital RT-PCR (RT-ddPCR) measurements of SARS-CoV-2 (N1 gene target) and Pepper Mild Mottle Virus (PMMoV) RNA in 40 wastewater RNA extracts. All three methods were highly sensitive, but appeared less accurate at very low concentrations. Lower inhibition was observed for RT-ddPCR than RT-qPCR with both SARS-CoV-2 and PMMoV targets, but inhibition appeared to be mitigated by dilution of template RNA. The concentrations of N1 and PMMoV from all three methods were significantly correlated (Pearson’s r=0.97-0.98 for N1 and r=0.89-0.93 for PMMoV), although RT-qPCR reported higher concentrations than digital methods. Taken together, this study provides support for the application of all three methods in wastewater-based epidemiology, with additional guidelines for the use of RT-qPCR.

**Impact Statement:** PCR-based assays are the current standard for sensitive, specific, rapid pathogen quantification in environmental samples, including wastewater. The increased availability of multiple digital PCR technologies necessitates side-by-side comparison between platforms, including traditional qPCR, to guide the application of these methods. Specifically, this work can inform interpretation of wastewater SARS-CoV-2 PCR data, as reported to public health agencies for pandemic response.

## 1. Introduction

SARS-CoV-2 wastewater-based epidemiology (WBE) is now widely applied as a means to monitor the spread of COVID-19 in communities (1,2). WBE may become more critical for providing accurate COVID-19 prevalence information as vaccines have become widely available in the U.S. and at-home testing has replaced clinical testing. WBE for human pathogens typically involves the extraction of nucleic acids from raw wastewater and PCR-based quantification of an RNA or DNA target. While a national database has been established for SARS-CoV-2 WBE in the United States (CDC-NWSS), there remains no standardized protocol, and laboratories are often limited by the equipment already available to them. Interlaboratory comparisons have shown one or more orders of magnitude differences in results reported by groups using different methods (3–5). While much work has focused on the comparison of viral concentration and RNA extraction methods from wastewater, there is limited data to specifically address differences introduced by the choice of quantification platform.

Three commonly-used reverse transcription polymerase chain reaction (RT-PCR) methods that have been applied for WBE are quantitative PCR (qPCR), fixed array-based digital PCR (dPCR), and droplet digital PCR (ddPCR). While qPCR is based on whole-sample real-time quantification relative to a standard curve, digital PCR methods rely on sample digitization or partitioning and use end-point detection to determine the number of positive microchambers or partitions. A Poisson distribution is used to calculate the initial concentration of the target in the sample based on the fraction of positive microchambers or partitions. Commonly cited advantages of dPCR and ddPCR include robustness to inhibition, high sensitivity, high reproducibility, and no need for a standard curve. However, droplet digital and digital PCR have somewhat more limited ability to measure high-concentration samples, meaning *a priori* knowledge is required for adequate dilution (5), and manual thresholding is sometimes required to determine separation between positive and negative droplets (for ddPCR) (6). While digital PCR is becoming more widespread, it is still out of reach for many laboratories due to the high initial cost of equipment and longer turnaround time from sample to results. Fixed array plate-based dPCR is technically more straightforward and faster to perform because it removes the droplet formation step, but it has lower throughput compared to ddPCR, and few studies have directly compared this technique to ddPCR and qPCR.

With the increased adoption of digital PCR platforms, there have been several extensive comparisons of ddPCR and qPCR applied for sensitive quantification of microorganisms in the environment (3–5). As SARS-CoV-2 WBE developed, many researchers performed methodological comparisons that included side-by-side tests of ddPCR and qPCR for SARS-CoV-2 gene targets (7–11). Overall, higher quantities were determined by qPCR than ddPCR, and, when measured, the correlation between results from the two platforms was reasonable (**Table S1**). However, conclusions were often limited by the small numbers of samples tested. Several investigations found that ddPCR was more sensitive and less likely to experience inhibition than qPCR (7,9,12,13), while one study suggested that inhibition of reverse transcription was greater in ddPCR than qPCR (11). In the most extensive of these studies, Ciesielski et al. (7) quantified SARS-CoV-2 in 63 wastewater samples with ddPCR and qPCR using the US CDC N2 assay. Fewer than half of the samples were above the defined limits of detection, and while inhibition was observed, the extent of inhibition in each platform was not determined. Just one study compared quantification of SARS-CoV-2 in wastewater using qPCR and fixed array-based dPCR (8). Notably, the N2 assay was previously found to be more sensitive in ddPCR (14), while the CDC N1 assay was more sensitive for qPCR (15,16).

Given the importance of accurate, reproducible WBE data despite differences in laboratory equipment, we sought to determine the comparability of three quantification methods–RT-qPCR, RT-ddPCR, and RT-dPCR—with RNA from northern California wastewater samples. We applied SARS-CoV-2 (CDC N1 diagnostic assay) and Pepper Mild Mottle Virus (PMMoV; fecal strength indicator (17–19)) assays on all three platforms. This study includes a direct comparison of 40 samples across three PCR platforms and a summary of the comparisons performed thus far in the literature. Sampling locations ranged from large treatment plants to residential buildings, allowing testing of a wide range of concentrations, sensitivity near the limit of detection, and the impact of inhibition.

## 2. Methods

### 2.1 Sample collection and RNA extraction

Forty raw wastewater samples were collected from locations across the San Francisco Bay Area between December 3rd and December 10th, 2020. In total, 30 unique locations were represented in the sample set, including wastewater treatment plant influents, subsewersheds, and residential buildings or campuses. For each sample, 40 mL of raw wastewater from a 24-hour composite sample was transferred to a 50 mL tube containing 9.35 g sodium chloride and 400 µL TE buffer (1 M TRIS, 100 mM EDTA). The sample tubes were shipped overnight with an ice pack to the University of California, Berkeley (UC Berkeley), and total nucleic acids were extracted within 48 hours of sampling. Nucleic acid extraction followed the 4S method (20). Briefly, the samples were heated at 70°C for 45 minutes, then filtered through 5 µm PVDF filters, mixed 1:1 with 70% vol/vol ethanol, bound to Zymo III-P columns (ZymoResearch), rinsed with wash buffers, and eluted with 200 µL ZymoPURE elution buffer. The resulting 200 µL of nucleic acid eluate for each sample was divided into four 50 µL aliquots and stored in LoBind tubes (Eppendorf) at -80°C.

Sample collection and extraction complied with the Environmental Microbiology Minimum Information (EMMI) guidelines (21). Collection was part of routine weekly monitoring conducted by the UC Berkeley COVID-WEB wastewater surveillance project. There was no evidence of contamination during sample collection in 10 months of sampling prior to this study. Therefore, environmental sampling controls were appropriately deemed unnecessary (21). Each sample was spiked with one of two amounts of the enveloped ssRNA virus Murine Hepatitis Virus (MHV, ATCC) (to a total spike amount of either 7.08 × 10^4^ gene copies or 7.08 × 10^5^ gene copies per 40 mL of raw wastewater) prior to heat inactivation. MHV served as a matrix recovery proxy to enable estimation of extraction efficiency of SARS-CoV-2. The post-extraction concentration was quantified using ddPCR to generate an extraction efficiency. A 40-mL phosphate buffered saline (1x PBS) negative extraction control was included with the batch of extractions on December 8th, 2020, and was quantified on one N1 plate and one PMMoV plate using RT-qPCR. No amplification of N1 was detected in the negative control and amplification of PMMoV was minimal (Cq=39.7).

### 2.2 RT-qPCR

After overnight storage at -80 °C, samples were thawed on ice and processed with RT-qPCR on a QuantStudio3 Real-Time PCR System qPCR machine (Thermo Fisher) at the University of California, Berkeley. The *One-Step RT-qPCR for SARS-CoV-2 Wastewater Surveillance protocol* (22) was followed. Briefly, each reaction contained 15 µL of reaction mix and 5 µL of template. The template consisted of undiluted RNA or five-fold diluted RNA in PCR water. The reaction mix contained TaqMan Fast Virus 1-Step Master Mix (Thermo Fisher Catalog #4444434) and primers and probes (Integrated DNA Technologies) in PCR water (see Greenwald et al. for sequences and concentrations (22). Methods complied with the Minimum Information for Publication of Quantitative Real-Time PCR Experiments (MIQE) guidelines (23).

The CDC N1 diagnostic assay was used for SARS-CoV-2 (24) and the coat protein gene was used for PMMoV (25). Both were singleplex assays with FAM-labeled probes. The N1 standard was made using the nucleocapsid (N) gene plasmid (Integrated DNA Technologies) grown in *E. coli*, digested to make a linearized plasmid stock, and quantified via Qubit, and Absolute Q (dPCR; see below). The PMMoV standard used on the first four plates of this study was an RNA ultramer (IDT), but it was found to have degraded (**Figure S1**). The standard used on the fifth plate was a linear DNA fragment (gBlock). For analysis, all PMMoV data were analyzed by applying the standard curve equation derived from the plate with the DNA standard (plate 1138; see Data Analysis).

Each of the six N1 and five PMMoV RT-qPCR plates contained samples, no-template controls (PCR water), and a standard curve, in triplicate wells (**Table S2**). All no-template controls were negative for all plates. The N1 standard plasmid was diluted in TE buffer to a stock concentration of approximately 1×10^7^ gene copies per microliter (gc/µL), later quantified by dPCR (see below). Serial dilutions in PCR water were performed to generate a seven point standard curve (5, 10, 20, 100, 1000, 10000, and 10000 gc/well, before adjusting based on the dPCR quantification of the standard). For N1 plates, standard curve efficiency ranged from 85% to 93% and R^2^ ranged from 0.90 to 1.0. A seven point standard curve was also used for PMMoV (10^2^ to 10^8^ gc/well). For PMMoV plates, the standard curve efficiency ranged from 84% to 122% and R^2^ ranged from 0.92 to 1.0. The concentration for each sample was determined by averaging the Cq values from all technical replicates that amplified and calculating a concentration based on the standard curve from each plate. Before averaging, outlier Cq values were removed from groups of technical triplicates using Grubb’s test (alpha = 0.05).

The limit of detection (LOD) for the N1 assay was determined to be 0.49 gc/uL RNA (2.43 gc per well), the point at which ≥ 95% of technical replicates were positive on 6 standard curves each run in triplicate (**Table S3**). The limit of detection for other assays was not investigated. Samples were called positive if at least 2 of 3 replicate wells amplified and the average concentration was above the LOD.

### 2.3 RT-dPCR

Extracted RNA samples were stored at -80 °C at UC Berkeley for one day past the last day of sampling, then hand-carried on dry ice to Combinati (Palo Alto, California) and stored at -80 °C for 50 additional days. Samples were thawed on ice and processed using RT-dPCR by Combinati. Steps complied with the digital MIQE guidelines (26,27). Each 9 µL reaction consisted of 1 µL of RNA, 2.25 µL of 4X Combinati 1-step RT-dPCR MasterMix, 0.45 µL of SARS-CoV-2 Wastewater Surveillance 4-plex assay (Combinati), and 5.3 µL water. Each plate contained negative controls (PCR water) and a positive control. The positive 4-target PCR control was made of synthetic single-stranded DNA containing the target N1, N2, and PMMoV sequences plus extracted Bovine Coronavirus RNA (PBS Animal Health).

After preparing the dPCR mix, 9 µL of the reaction mixture was loaded into the MAP16 plate followed by an overlay of 15 µL of isolation buffer. The prepared MAP16 plate was then loaded on the Absolute Q instrument (Applied Biosystems QuantStudio Absolute Q Digital PCR system). Thermocycling was performed using Absolute Q with the following program: reverse transcription at 50 °C for 10 minutes, preheating (enzyme activation) at 95 °C for 10 minutes, and 45 cycles of denaturation at 95 °C for 5 seconds and annealing/extension at 55 °C for 30 seconds.

The limit of detection and limit of quantification used for the assay were both two positive microchambers per reaction (∼0.2 gc/µL). These values are the same because digital PCR provides absolute quantification without the need for a standard curve. Data analysis was performed on the Absolute Q Analysis Software (v4.2.1), which reports the concentration of each reaction in gene copies per microliter (gc/µL) (**Table S4**). The initial sample concentration was manually calculated by adjusting the reported dPCR concentration based on the input sample volume for each sample. The arithmetic mean concentration of the duplicate measurements was calculated and reported for each sample. Samples were determined to be positive if at least 3 total microchambers were positive across duplicate reactions.

### 2.4 RT-ddPCR

Extracted RNA samples were stored at -80 °C at UC Berkeley for 41 days past the last day of sampling, then hand-carried on dry ice to Bio-Rad (Pleasanton, California) and stored at -80 °C for two additional days. Samples were thawed on ice and processed using the One-Step RT-ddPCR Advanced Kit for Probes (Bio-Rad); 9 µL was used for a 20 µL reaction volume and samples were analyzed without technical replicates. Each droplet was 0.85 nL. Steps complied with the digital MIQE guidelines (26,27). The primer and probe concentration in the reaction was 900 nM and 250 nM, respectively. Assays were purchased from IDT and manufactured under SARS-CoV-2 template-free conditions. The CDC N1 target for SARS-CoV-2 (HEX-labeled probe) (24) and PMMoV target (FAM-labeled probe) were run in singleplex reactions. The CDC N2 (24), E (28), and MHV targets (FAM-, HEX- and FAM-mixture, and HEX-labeled probes, respectively) were run as a triplex reaction (**Table S5**). Each reaction was dropletized using the QX200 Auto DG (Bio-Rad). Thermal cycling was performed using a C1000 Touch (Bio-Rad) with the following program: reverse transcription at 50 °C for 60 minutes, enzyme activation at 95 °C for 10 minutes, 40 cycles of denaturation at 94 °C for 30 seconds and annealing/extension at 55 °C for 1 minute, enzyme deactivation at 98 °C for 10 minutes, and a final hold at 4 °C. The ramp rate was set to 2 °C/second.

Samples were analyzed without dilution and at a 1:5 dilution (7 µL diluted into a final volume of 35 µl with nuclease free water). Synthetic SARS-CoV-2 RNA (Exact Diagnostics) was used as a positive control on all runs. Nuclease free water was used as a no-template control on all runs (1-2 wells per run) and all negative controls were negative.

Data were analyzed using the QXManager v1.2. All wells were thresholded manually and were confirmed to have appropriate droplet counts (**Table S6**). A sample was interpreted as positive if the ddPCR concentration for N1 was ≥0.1 gc/µL (RNA sample concentration of 0.22 gc/µL) and there were two or more positive droplets per sample (no technical replicates were performed). This limit of detection was determined using a serially diluted heat-inactivated virus to establish the lowest detectable concentration of SARS-CoV-2 at which at least 95% of true positive replicates tested positive.

### 2.5 Quantification of qPCR standards by dPCR

Two aliquots of the RT-qPCR standards for N1 and PMMoV were hand carried on dry ice to Combinati for quantification. The standards were diluted 1000-fold, 10,000-fold, and 100,000-fold before quantification using the Absolute Q digital PCR platform. Each dPCR reaction consisted of 1 µL of diluted standard material, 2.25 µL of 4X Combinati 1-step RT-dPCR MasterMix, 0.45 µL of 4-plex Wastewater assay, and 5.3 µL water. Each dilution point was quantified in duplicate. After correcting the reported concentration based on the appropriate dilution factor, the absolute concentration of the standard material was calculated using the average values of both tubes across the dilution series (**Table S7**). The N1 (linearized plasmid) and PMMoV (dsDNA gBlock) standard concentrations used in data analysis were based on quantification of the standards via Absolute Q dPCR (**Table S2**).

### 2.6 Data Analysis

Data analysis was performed using Python (v3.9.5) using modules Pandas (v1.2.5), NumPy (v1.21.0), SciPy (v1.7.0), and Plotnine (v0.8.0). RT-qPCR analysis of standard curves and unknowns was performed using custom code as previously described (29) (https://github.com/wastewaterlab/data_analysis). Pearson correlation coefficients were determined using SciPy, and linear regressions were conducted using NumPy polyfit. Paired concentrations of diluted and undiluted samples were compared using the Mann-Whitney U test (SciPy) to determine whether there was significant inhibition during RT-PCR. All analysis code can be found at https://github.com/wastewaterlab/data_analysis/blob/master/notebooks/pcr_comparison.ipynb.

## 3. Results

A total of 40 raw wastewater samples from 30 distinct residential, subsewershed, and treatment plant influent locations in the San Francisco Bay Area underwent large-volume nucleic acid extraction at UC Berkeley. Quantification of SARS-CoV-2 (CDC N1 diagnostic assay) and PMMoV (coat protein gene) RNA was performed by three laboratories using qPCR, dPCR, and ddPCR, respectively (**Table S8**). Undiluted RNA was quantified for all samples, while five-fold diluted RNA was also quantified with qPCR and ddPCR to assess inhibition. All three methods produced similar trends across samples that ranged three orders of magnitude in concentration (**Figures 1 and S2**).

**Figure 1.**
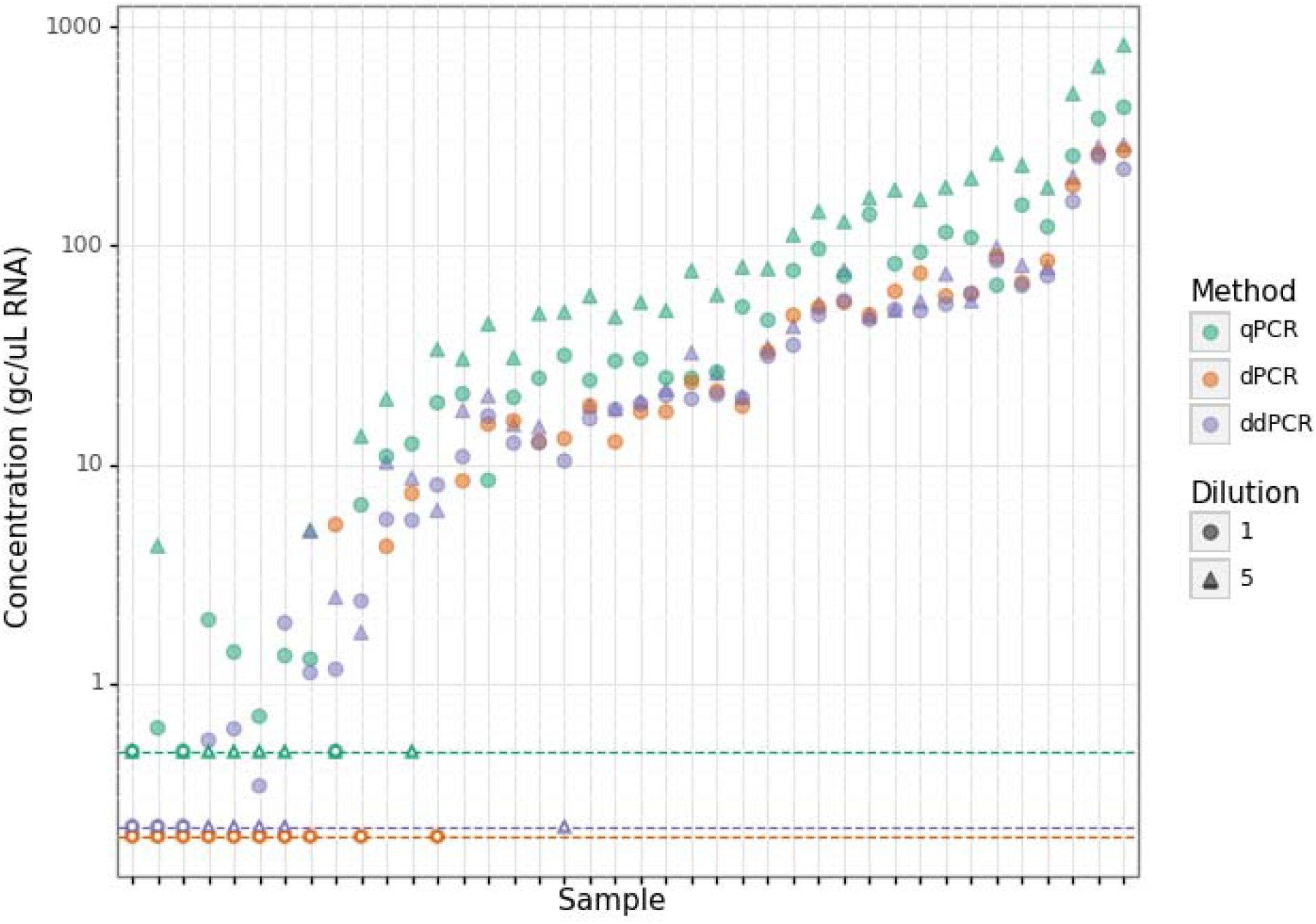
Comparison of SARS-CoV-2 and PMMoV concentrations in RNA from 40 wastewater samples from the San Francisco Bay Area. Samples are ordered by average N1 concentration rank for each method across all methods. Each sample was measured undiluted via dPCR and both undiluted and five-fold diluted via ddPCR and qPCR. The N1 limit of detection is shown for each method with a dashed line; all non-detects are plotted as open points at the LOD for each method. Concentrations shown for diluted qPCR and ddPCR samples are corrected for the dilution factor.

### 3.1 Sensitivity

The limits of detection for the N1 assay with qPCR, dPCR and ddPCR were found to be 0.5, 0.2, and 0.22 gene copies per microliter (gc/µL) RNA, respectively (see Methods). The sensitivity of each platform is also impacted by PCR inhibition (see below) and by the volume of template RNA included in the PCR reaction. For a single reaction well, qPCR used 5 µL, dPCR used 1 µL, and ddPCR used 9 µL. Additionally, measuring the same sample in technical replicate wells may allow for improved sensitivity. Here, qPCR utilized 3 wells, dPCR utilized 2 wells, and ddPCR utilized 1 well (no replicates). Of 40 samples compared with the N1 assay (undiluted), there was total agreement on 2 negatives and 29 positives across all platforms (**Table S8**). ddPCR and dPCR jointly reported 1 additional negative, qPCR reported 1 additional negative, and dPCR reported 7 unique negatives (**Figure 2**). All but one of the negatives were samples from residential sewersheds (buildings or campuses). Notably, the Murine Hepatitis Virus (MHV) matrix recovery control had >50% calculated recovery efficiency for all but five samples. Of those five, two were non-detects with N1 in dPCR only. This suggests that, overall, non-detects were not due to poor extractions.

**Figure 2.**
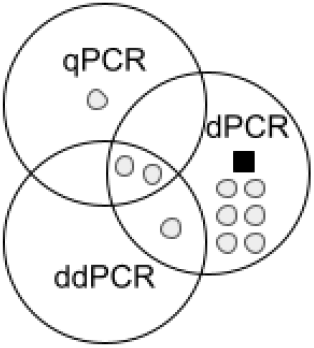
Venn diagram of the samples with non-detects in one or more PCR method. Samples are depicted as squares (a single WWTP influent site) or circles (residential sites).

### 3.2 Inhibition

Next, we assessed inhibition, which can lead to under-quantification of the molecular target or false negative results (15,30). Inhibition is especially a concern in WBE because of the inhibitory substances found in wastewater and because inhibition impedes comparison between laboratories that use different molecular methods (31). To test for inhibition, we compared the results of qPCR and ddPCR with and without prior dilution of the RNA template (21,32). If inhibitors were present, dilution would decrease their concentration in the PCR reaction, resulting in higher reported concentrations after accounting for the dilution factor. Five-fold dilution was chosen after prior qPCR testing of 5-, and 10-fold dilutions revealed that 10-fold often led to loss of signal (17). We quantified N1 and PMMoV in undiluted and five-fold diluted samples using qPCR and ddPCR (**Figure 1**) and calculated the ratio between diluted and undiluted concentrations (**Figure 3**). Median diluted-to-undiluted ratios for ddPCR and qPCR were 1.15 vs. 1.82 for N1 and 1.17 vs. 1.88 for PMMoV, indicating that qPCR results were significantly more affected by inhibition (two-sided Mann-Whitney U test p=1.2×10^−8^ for n=31 comparisons of N1 and p=3.6×10^−10^ for n=33 comparisons of PMMoV).

**Figure 3.**
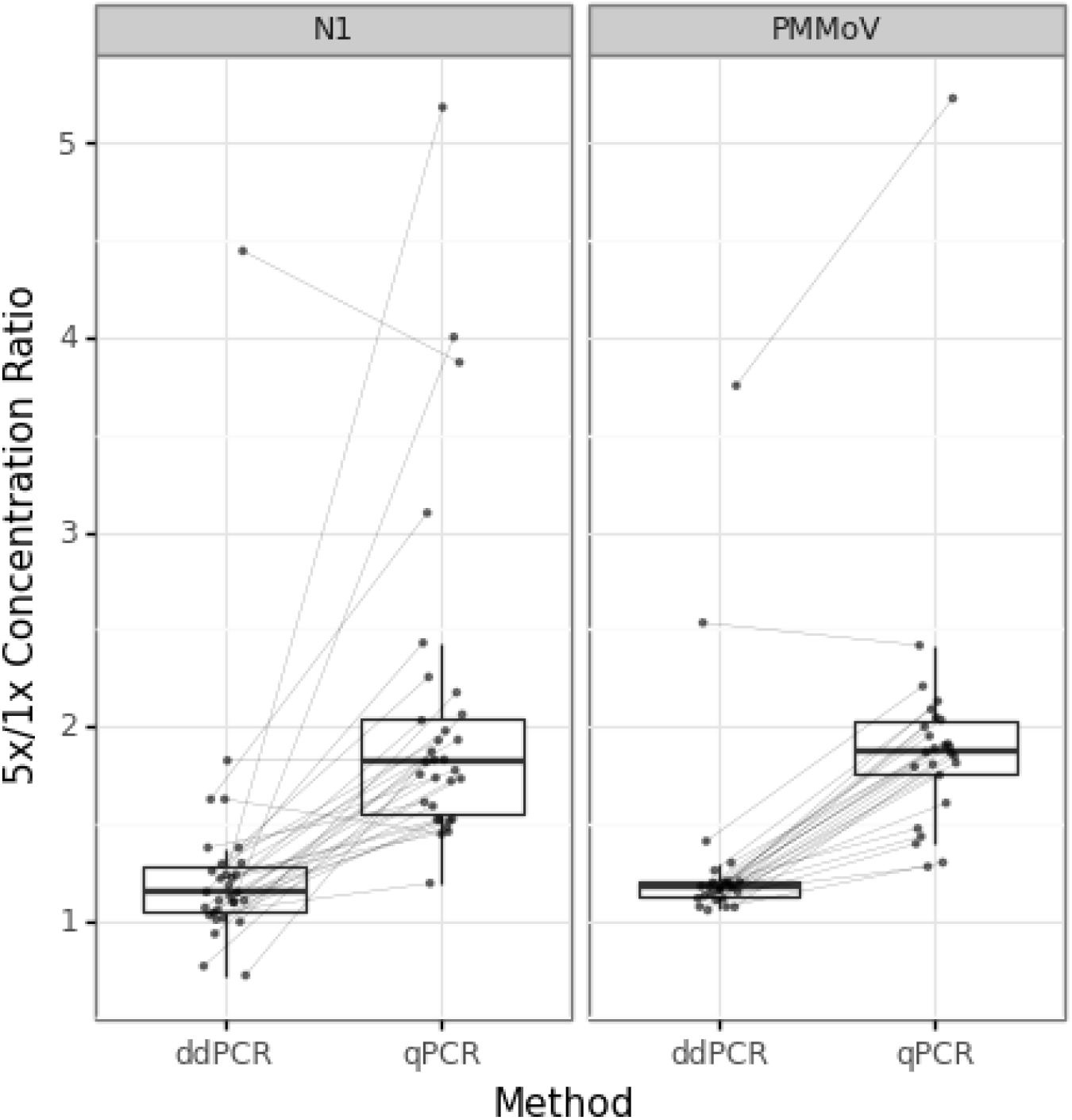
Ratio of target quantification in five-fold diluted sample relative to undiluted sample for ddPCR and qPCR. The concentrations for the diluted samples were corrected for the dilution factor (multiplied b 5) prior to calculating the ratio. Samples with non-detects are not shown. The boxes show the first and third quartiles and the whiskers extend to the largest value within 1.5 times the interquartile range. Individual data points are shown in gray and samples are connected by gray lines.

### 3.3 Quantification

We found that N1 and PMMoV results from all three methods were linearly correlated after log10 scaling with significant Pearson correlation coefficients for each pair of methods (**Table 1, Figures S3 and S4**). Correlations were determined using the data from 5-fold dilutions of qPCR and ddPCR due to the presence of inhibition in some samples (see above). Samples that were non-detect with any method were removed prior to calculation. Despite the strong correlation, undiluted qPCR concentrations were higher than dPCR concentrations (median fold difference = 1.7) and ddPCR concentrations (median fold difference = 1.8). Notably, qPCR standards were quantified by dPCR, which reported concentrations for N1 and PMMoV that were, respectively, 60.8% and 39.2% lower than those found with Qubit (**Table S7**). This difference was accounted for during data analysis.

**Table 1.** Correlation statistics and linear regressions for log10-scaled data from each pair of PCR methods.

| Target | Independent | Dependent | n | Slope | Intercept | Pearson's r* | Spearman r* |
| --- | --- | --- | --- | --- | --- | --- | --- |
| N1 | qPCR | dPCR | 27 | 1.06 | -0.60 | 0.98 | 0.97 |
|  | qPCR | ddPCR | 27 | 0.93 | -0.27 | 0.97 | 0.95 |
|  | dPCR | qPCR | 27 | 0.90 | 0.63 | 0.98 | 0.97 |
|  | dPCR | ddPCR | 27 | 0.86 | 0.29 | 0.97 | 0.97 |
|  | ddPCR | qPCR | 27 | 1.02 | 0.38 | 0.97 | 0.95 |
|  | ddPCR | dPCR | 27 | 1.11 | -0.24 | 0.97 | 0.97 |
| PMMoV | qPCR | dPCR | 40 | 1.04 | -0.46 | 0.90 | 0.90 |
|  | qPCR | ddPCR | 40 | 1.03 | -0.28 | 0.93 | 0.91 |
|  | dPCR | qPCR | 40 | 0.78 | 1.13 | 0.90 | 0.90 |
|  | dPCR | ddPCR | 40 | 0.85 | 0.68 | 0.89 | 0.94 |
|  | ddPCR | qPCR | 40 | 0.84 | 0.81 | 0.93 | 0.91 |
|  | ddPCR | dPCR | 40 | 0.93 | 0.16 | 0.89 | 0.94 |

\* All correlations were significant ( $p < 1.5 \times 10^{-12}$ )

## 4. Discussion

All three PCR methods produced quantitative measurements of N1 and PMMoV RNA concentrations across three orders of magnitude (**Figure 1**), and concentrations were strongly correlated between methods (**Table 1**). Despite quantification of the qPCR standards with dPCR prior to analysis, measured qPCR concentrations were consistently higher than dPCR and ddPCR concentrations, which were in closer agreement. This trend has also been observed in previous studies (**Table S1**), indicating a need to normalize concentrations before comparisons can be made between measurements using different methods. One possible explanation for this discrepancy is that qPCR standard curves may be affected by DNA adherence to the walls of plastic tubing or degradation of DNA samples, leading to overestimation of the DNA standard, and subsequent over-quantification of the wastewater sample. Conversely, these same factors should affect the sample nucleic acids as well, resulting in potential under-quantification of samples by ddPCR and dPCR. Another possible explanation is that the reverse transcription step in RT-qPCR could have produced more than one cDNA molecule per RNA template, resulting in over-quantification of the sample material (RNA) but not the DNA standards. Meanwhile, the reverse transcription step in digital PCR methods was conducted within the partitions/microchambers, and a proportional increase in cDNA would not affect the readout of these endpoint PCR assays.

Also consistent with prior work (**Table S1**), inhibition was significantly lower in ddPCR than qPCR based on the lower diluted-to-undiluted ratio in ddPCR (**Figure 3**). Nonetheless, inhibition in qPCR does not appear to have led to many false negatives relative to digital PCR methods (**Figure 2**). Additionally, while inhibition was not measured with dPCR, similar results would be expected given that both digital PCR methods measure the presence or absence of DNA template in individual microchambers and are therefore less impacted by the decrease in amplification efficiency caused by PCR inhibitors (12,33).

Lastly, limits of detection measurements are a function of the total sample volume included in the reaction and for digital PCR, the number partitions or microchambers measured, but these are not standardized across platforms. Additionally, the method for defining the limit of detection was different for each platform. Thus, conclusions comparing the sensitivity of each platform based on the defined LoDs should be taken with care.

## 5. Conclusion

All three methods are highly sensitive and quantitative, making them appropriate tools for use in WBE. However, based on our findings, additional guidelines for RT-qPCR should be followed when this method is used for WBE. First, to increase the accuracy of RT-qPCR, dilutions should be performed when there are signs of inhibition. Undiluted samples should also be run in parallel in case dilution causes the signal to drop below detectable levels. The same should be performed for digital PCR, if sample volumes and logistical constraints allow. Second, the differences between Qubit and fixed array-based digital PCR measurements of the DNA standard in this study (see Methods) point to the importance of the method of quantification for qPCR standards. We suggest that WBE databases such as CDC-NWSS should track how qPCR standards are quantified, and this should be reported in all qPCR-based WBE studies. For internal consistency, long-term monitoring projects should limit changes to standards and carefully compare new batches of standards as part of quality control.

Third, we have found that results of long term-monitoring using RT-qPCR may be subject to plate-to-plate fluctuations due to differences in standard curve serial dilutions (29). While standard curve efficiency and R^2^ are key components of quality control, the y-intercept should also be monitored as an indicator of standard degradation or batch differences. Finally, we note that all methods are less accurate at low concentrations due to stochasticity (whether the RNA molecules are captured in the subsample that is taken for PCR analysis). Here, testing more technical replicates may improve accuracy, and replication may be most important for presence/absence determinations in residential facilities. We also expect that including more RNA template in the PCR reaction would improve accuracy. The sample volume included in the PCR reactions was a major difference between the platforms, as ddPCR utilized 9 µL, RR-qPCR utilized 5 µL, and dPCR utilized 1 µL. This may explain why SARS-CoV-2 was detected in more samples using RT-qPCR and ddPCR compared to dPCR.

Digital PCR methods are advantageous for their ability to rapidly bring online multiplexed assays for new SARS-CoV-2 mutation detection or other pathogens. Additionally, the absolute quantification and increased tolerance to inhibition of dPCR and ddPCR allows higher confidence in the quantitative comparison of assays for multiple targets from the same organism (e.g. to calculate the percentage of total SARS-CoV-2 corresponding to a variant lineage). Despite these advantages, qPCR is a familiar, more widely available method for many researchers globally, and it can be fast, cost-effective, and high-throughput. Overall, the data presented in this study and summarized from previous work provide evidence that, when used appropriately, qPCR, dPCR, and ddPCR are all suitable WBE methods that generate highly correlated results for a wide range of wastewater sources and target concentrations.

## Data Availability

All data produced in the study are available in the supplementary tables

## 5. Acknowledgements

We thank Christina Bouwens and Robert Lin (Thermo Fisher) for their collaboration to run samples on the Absolute Q. We thank our partners at the following agencies and facilities for sample collection: Sanitary District No. 5 of Marin County; Central Marin Sanitation Agency; East Bay Municipal Utility District; San Francisco Public Utilities Commission; Las Gallinas Sanitary District; Sewerage Agency of Southern Marin; Novato Sanitary District; Central Contra Costa Sanitary District; Delta Diablo; West County Water District; Yountville; American Canyon; California Medical Facility; and University of California, Berkeley. We also thank volunteers in the UC Berkeley COVID-WEB wastewater testing laboratory. We gratefully acknowledge funding from the Catena Foundation. A.W.H., H.D.G., and L.C.K. were supported by the National Science Foundation (NSF) Graduate Research Fellowship [grant number DGE-1752814].

