## supplementary figures for "Comparison of RT-qPCR and Digital PCR Methods for Wastewater-Based Testing of SARS-CoV-2"

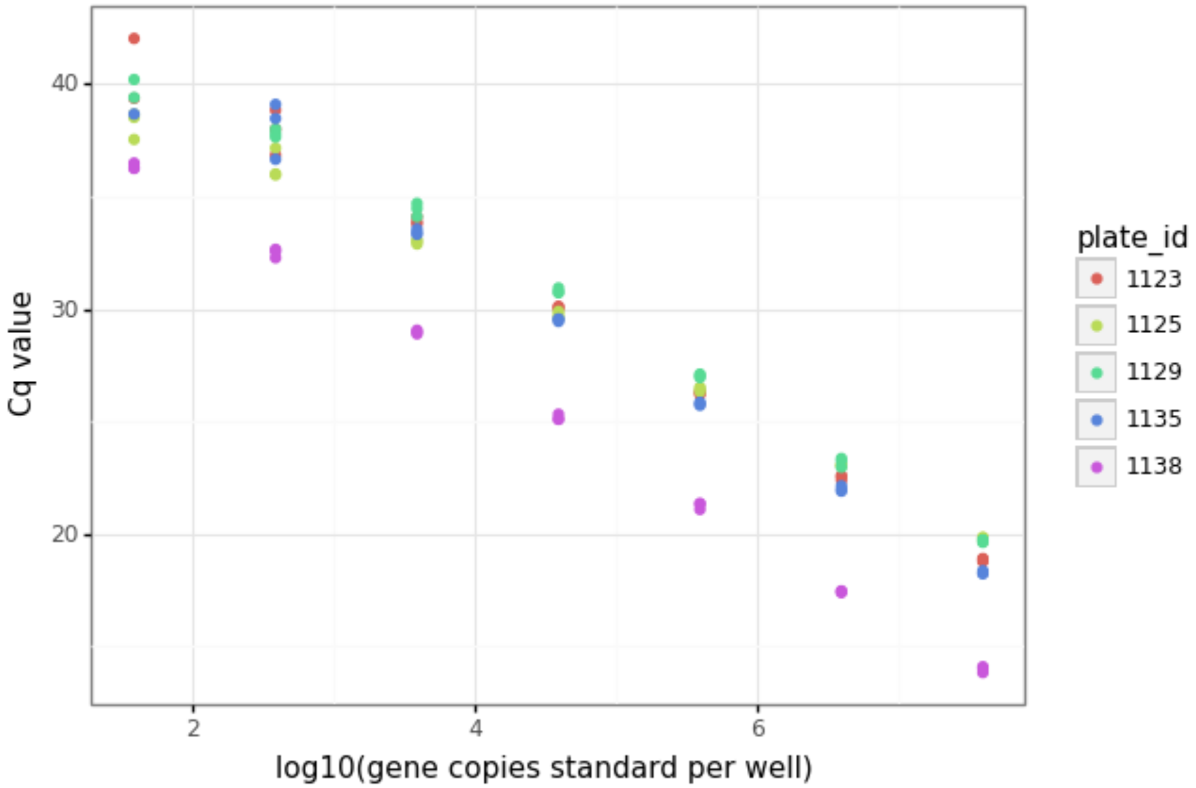


**Figure S1.** RT-qPCR standard curves for five PMMoV assay plates. Plates 1123, 1125, 1129, and 1135 used an RNA ultramer standard that had degraded, while plate 1138 used a fresh dsDNA gBlock standard that was quantified via ddPCR (see Table S7). All PMMoV data were analyzed using the standard curve from plate 1138.

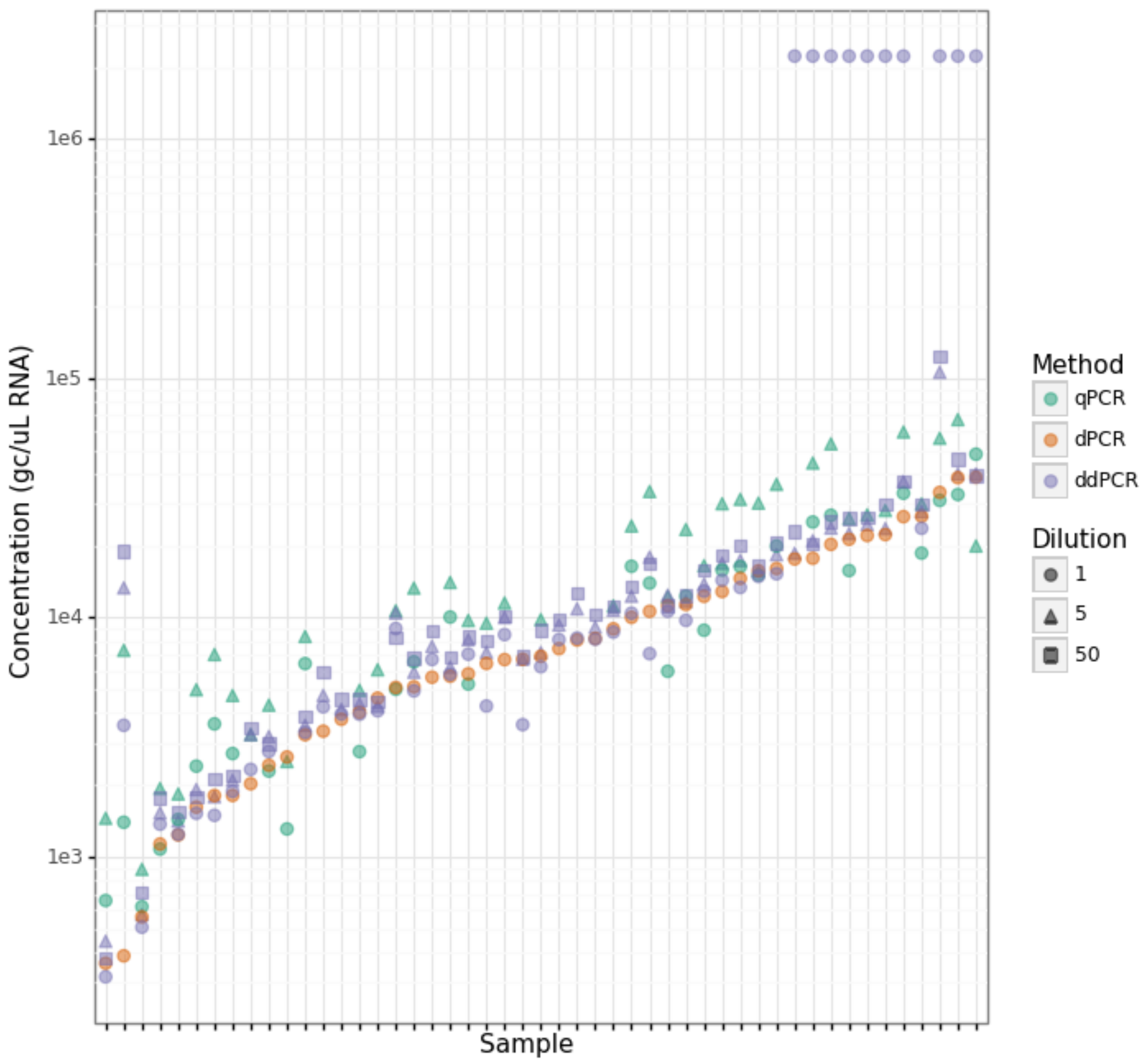


**Figure S2.** PMMoV concentrations of wastewater-derived RNA determined by the three PCR methods, ordered by concentration in dPCR. RT-qPCR was performed on 5-fold diluted RNA and, for some samples, also performed on undiluted RNA. RT-ddPCR was performed on undiluted, 5-fold diluted, and 50-fold diluted RNA. Where all partitions were positive, ddPCR data were substituted with $2.2 \times 10^6$ gc/uL RT-dPCR was performed on undiluted RNA only.

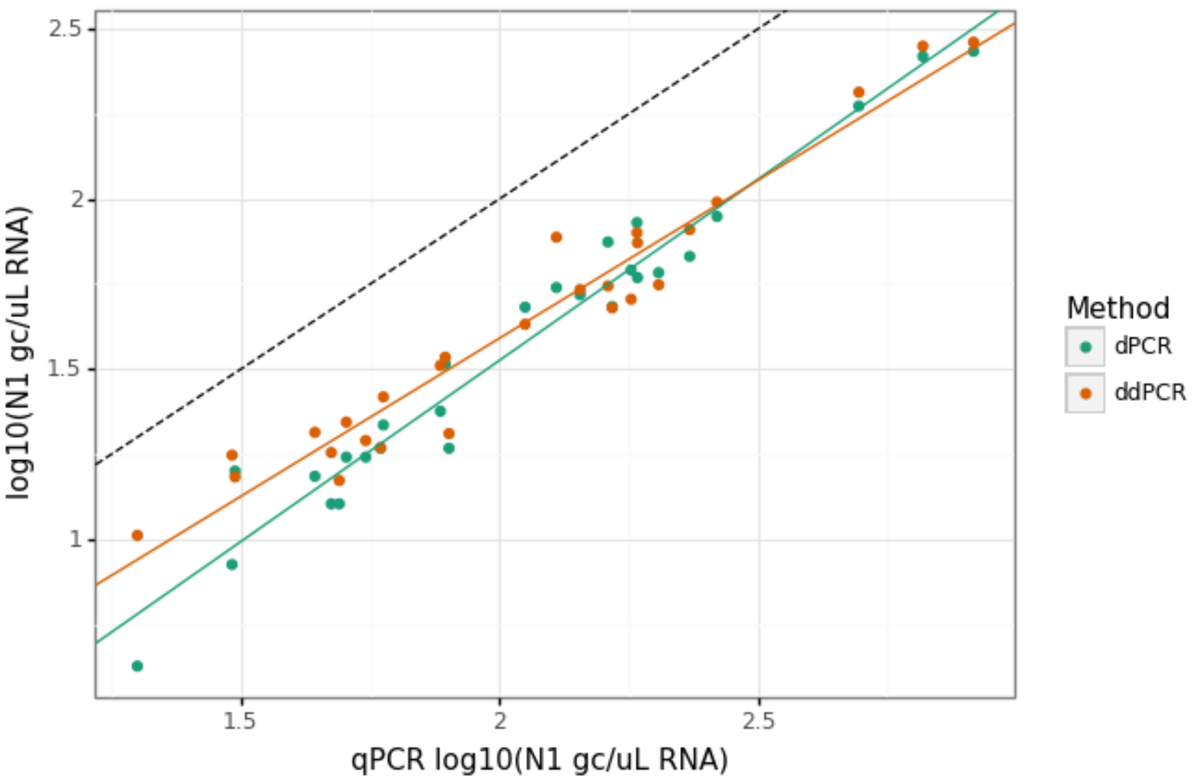


**Figure S3.** Linear regression of log10-scaled quantification of the N1 target via dPCR (x-axis) versus qPCR and ddPCR (y-axis).

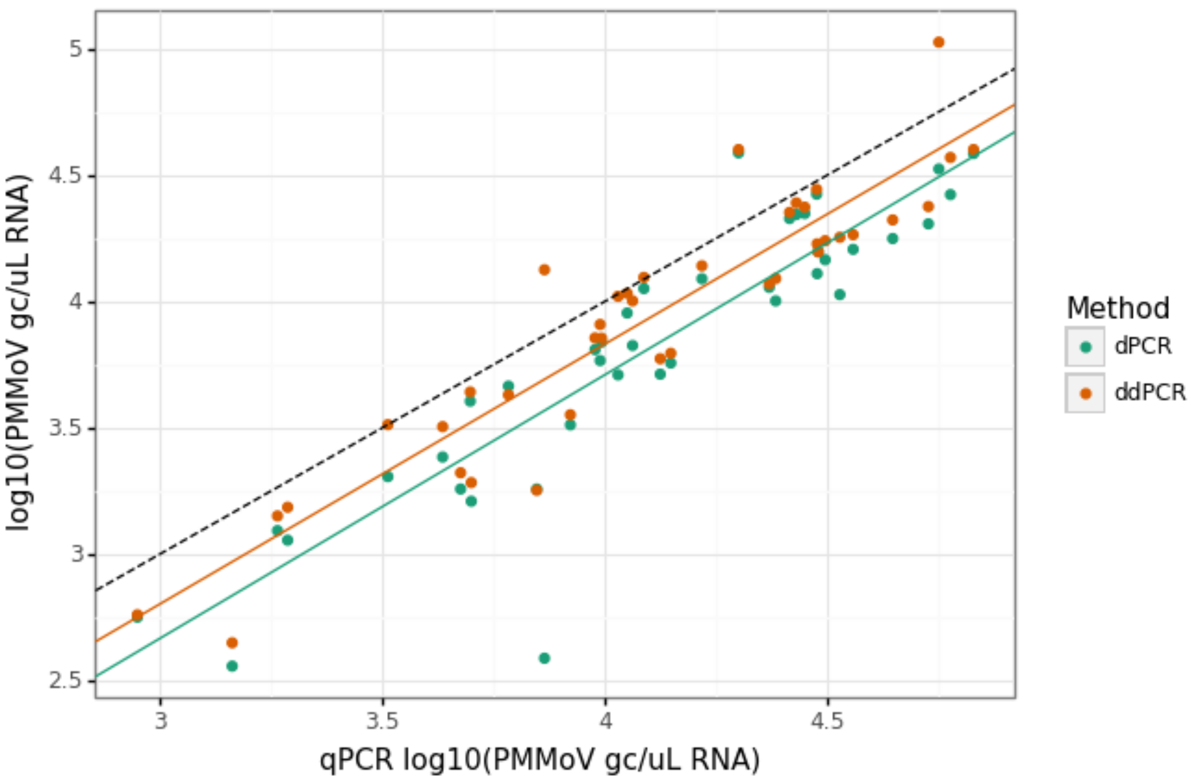


**Figure S4.** Linear regression of log10-scaled quantification of the PMMoV target via dPCR (x-axis) versus qPCR and ddPCR (y-axis).
